# NEW CORONAVIRUS IN PREGNANT WOMEN. Maternal and perinatal outcomes

**DOI:** 10.1101/2021.06.03.21258328

**Authors:** Jaime Sánchez, Jorge Espinosa, Luis Carlos Caballero, B Sara Campana, Arelys Quintero, Carlos Luo, C Jorge Ng, Rafael de Gracia, Paulino Vigil-De Gracia

**Affiliations:** Ginecología y Obstetricia, Hospital Santo Tomás, Panamá, Panamá; Ginecología y Obstetricia, Hospital Luis “Chicho Fábrega”, Santiago-Veraguas, Panamá; Ginecología y Obstetricia, Complejo Hospitalario Dr. AAM CSS”, Panamá, Panamá; Ginecología y Obstetricia, Hospital José Domingo de Obaldía, David-Chiriquí, Panamá; Ginecología y Obstetricia, Hospital San Miguel Arcangel, Panamá, Panamá

**Keywords:** SARS-CoV-2 infection, pregnancy, premature rupture of membranes, maternal infection, neonatal complications, maternal lethality

## Abstract

**Objectives:** To report the maternal and neonatal results of patients infected with COVID-19 in Panama.

**Methods:** The study is based on the analysis of pregnant women with COVID-19, in 5 hospitals in the Republic of Panama. The inclusion criteria were: Patients with or without symptoms, positive RT-PCR for SARS-CoV-2 in the period from March 23 to 6 months after, whose births were attended in one of those 5 hospitals and who signed the consent. Data was obtained at the time of diagnosis of the infection and at the time of termination of pregnancy for the mother and newborn.

**Results:** 253 patients met the inclusion criteria. Most were diagnosed in the third trimester (89.3%). 10.3% of the patients presented in a severe form of COVID-19. The most frequent complication was pre-eclampsia and if we add gestational hypertension they represent 21.2%; most of the patients terminated the pregnancy by caesarean section (58%). 26.9% (95% CI 21.3-32.9%) of the births were premature, and perinatal mortality was 5.4% (95% CI 3.0-9.0%). There was a need for mechanical ventilation in 5.9% (95% CI 3.6-9.6%) of the cohort and there were four maternal deaths (1.6% - 95% CI 0.6-4.0%).

**Conclusions:** This study of pregnant women infected with COVID-19 and diagnosed with RT-PCR shows serious maternal complications such as high admission to the ICU, need for mechanical ventilation and one death in every 64 infected. Frequent obstetric complications such as hypertension, premature rupture of membranes, high rate of prematurity and perinatal lethality were also seen.

## INTRODUCTION

Since the end of 2019 it has been detected in Wuhan-China, a viral infection whose causative agent was a new coronavirus, later identified as SARS-CoV-2; this viral disease led the World Health Organization to declare the COVID-19 pandemic for March 11 ^1-3^.

One year after the appearance of this disease, much progress has been made in the knowledge of its epidemiological behavior, virology, genetics, clinical findings, lethality and affectation by age groups and baseline conditions of the population ^4^. We have learned during this first year about the associated complications, risk of hospitalization, greater possibility of admission to an intensive care unit, greater possibility of mechanical ventilation and greater possibility of death ^5^, in pregnant women with COVID-19. Furthermore, we know that its evolution is less severe than other respiratory diseases caused by coronavirus such as SARS and MERS ^6^. High frequencies of cesarean sections, premature births, ruptures of membranes and other complications have been associated with COVID-19 ^7^. From the neonatal side, prematurity, low weight and pneumonia are the most frequent findings ^5-7^, vertical transmission has not been proved with certainty ^8^ and perinatal transmission is slightly more frequent but its impact on neonatal health is mild ^9^.

As time passes, more reports are made about the new coronavirus and pregnancy, but few reports are from third world countries and Latin America. It is necessary to know about the evolution and results that the SARS-CoV-2 infection produces around the world and thus have a more accurate knowledge of its global impact. Our objective is to report the findings and evolution of pregnant women with SARS-CoV-2 in 5 main hospitals in the Republic of Panama during the first 6 months of the epidemic.

## MATERIALS AND METHODS

We conducted a prospective and observational cohort study from the beginning of the epidemic in Panama (mid-March 2020) for a period of 6 months, in pregnant women with a diagnosis of SARS-CoV-2 confirmed by RT-PCR. Pregnant women with or without symptoms could be part of the study. Informed consent was required and the patients had to be admitted or delivered in one of the following hospitals: Complejo Hospitalario “Dr. AAM “Social Security Fund in Panama, Santo Tomás Hospital in Panama, José Domingo de Obaldía Hospital in Chiriquí, Luis” Chico “Fábrega Hospital in Veraguas and San Miguel Arcángel Hospital in Panama.

The sampling, processing, and laboratory techniques in patients with clinical suspicion, contacts, or specific indication were done following the national guidelines given by the ministry of health of Panama, which are those required by the World Health Organization.

Maternal data such as age, parity, symptoms, body mass index, comorbidities, gestational age at the onset of symptoms and at the end of pregnancy, severity of the disease, admission to the intensive care unit, need for mechanical ventilation and evolution were recorded. In addition, obstetric complications, cause of pregnancy termination, birth route, birth weight, APGAR at the first and fifth minute, time between the onset of symptoms and the termination of pregnancy were recorded.

The protocol was approved by the national bioethics committee of Panama, protocol reference: EC-CNBI-2020-04-45.

The maternal baseline characteristics and the neonatal results are presented in absolute numbers, percentages with 95% CI, means and standard deviation.

In view of incomplete data in some cases, absolute numbers were paired with total number of cases in which the information about the characteristic being studied was available. Statistical analysis was performed with Epi Info software version 7 (Centers for Disease Control and Prevention, Atlanta, GA).

## RESULTS

Since March 23, when the first confirmed pregnant case was registered in Panama, up to 6 months later, 253 cases with positive SARS-CoV-2 by RT-PCR were registered (in the five hospitals), eleven (4.35%) were abortions, 21 have not finished the pregnancy yet or were treated in other hospitals after their recovery. Maternal and birth data were obtained in 221 patients. Most participants started symptoms or were diagnosed in the third trimester (89.3%), see more information in table1. 56.9% (95% CI 50.8-62.9%) of the patients presented symptoms at the time of diagnosis, fever being the most common symptom, but in only 36% (95% CI 30.3-42.1%) of the cases; 10.3% (95% CI 7.1-14.6%) of the patients presented with severe disease, see table 1.

**TABLE 1.**
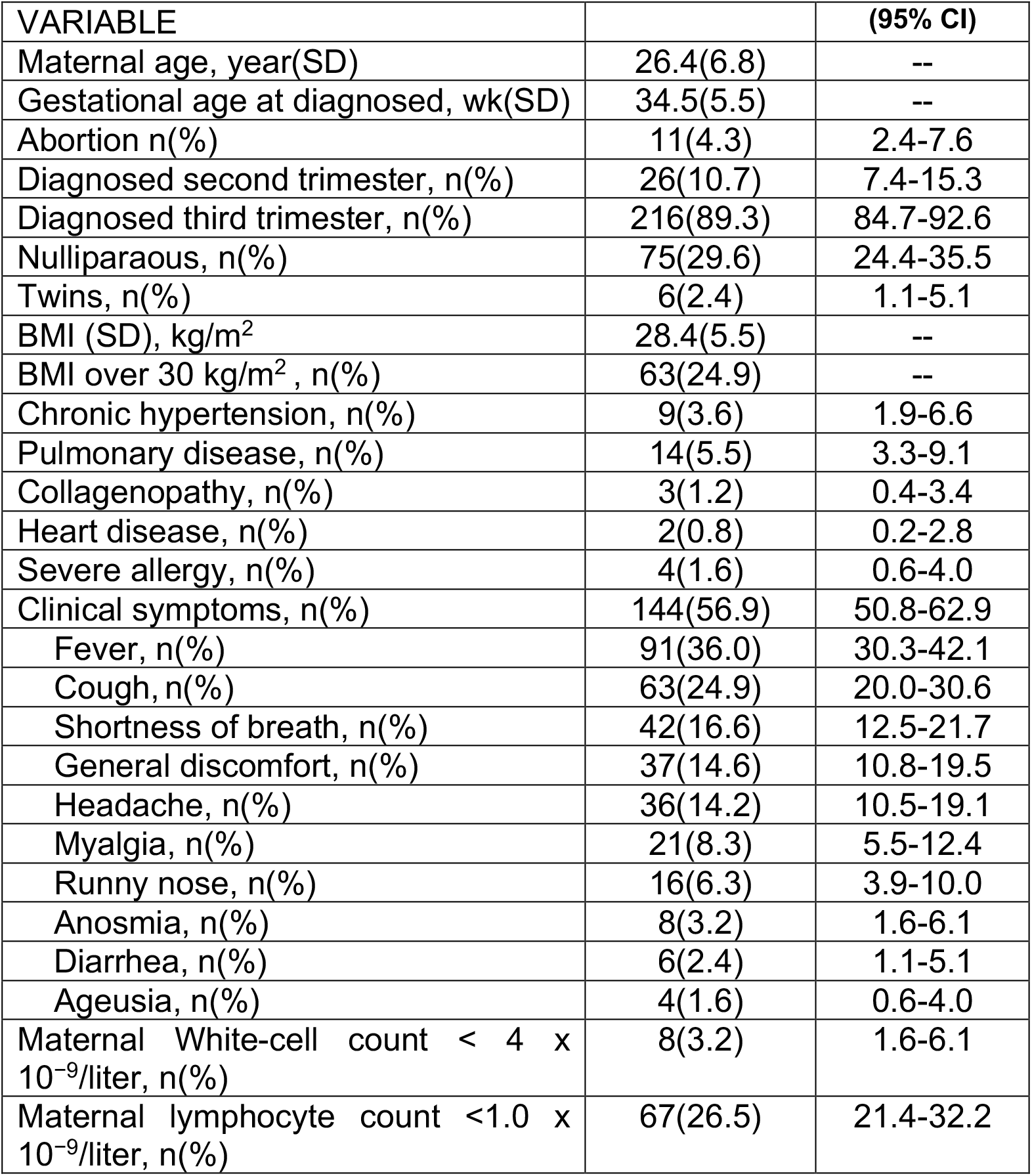
Demographic, clinical and laboratories characteristics of pregnant women infected by SARS-CoV-2.

The most frequent complication was pre-eclampsia and if we add gestational hypertension they represent 21.2%; most of the patients ended the pregnancy by cesarean section (58%), however, in only 10.1% the indication was the disease itself (COVID-19), Table 2.

**TABLE 2.** Therapy prescribed and maternal outcomes to pregnant women infected by SARS-CoV-2

|  |  |  |
| --- | --- | --- |
| Mother antibiotics, n(%) | 38(15.0) | 11.1-19.9 |
| Mother hydroxychloroquine, n(%) | 10(4.0) | 2.2-7.1 |
| Mother lopinavir–ritonavir, n(%) | 5(2.0) | 0.9-4.6 |
| Mild forms, n(%) | 227(89.7) | 85.5-92.9 |
| Severe forms, n(%) | 26(10.3) | 7.1-14.6 |
| Mother Intensive care unit admission, n(%) | 17(6.7) | 4.2-10.5 |
| Mother Endotracheal intubation, n(%) | 15(5.9) | 3.6-9.6 |
| Maternal death, n(%) | 4(1.6) | 0.6-4.0 |

**TABLE 3.** Obstetrics and clinical outcomes of neonates born from women infected by SARS-CoV-2 *.

|  |  | (95% CI) |
| --- | --- | --- |
| Gestational age at delivery, wk(SD) | 37.2(3.3) | -- |
| Pre-eclampsia, n(%) | 34(15.4) | 11.2-20.7 |
| Gestational hypertension, n(%) | 13(5.9) | 3.5-9.8 |
| Gestational diabetes, n(%) | 6(2.7) | 1.3-5.8 |
| Normal vaginal delivery, n(%) | 93(42.1) | 35.8-48.7 |
| Cesarean delivery, n(%) | 128(57.9) | 51.3-64.2 |
| Indicated by COVID-19, n(%) | 13(10.2) | 6.0-16.6 |
| Preterm delivery < 37 weeks, n(%) | 59(26.7) | 21.3-32.9 |
| Small-for-gestational age/fetal-growth restriction, n(%) | 27(12.2) | 8.5-17.2 |
| Premature rupture of membrane, n(%) | 18(8.1) | 5.2-12.5 |
| Birth weight g (SD) | 2894.0(769.4) | -- |
| APGAR 1 minute (IQR) | 8(8-9) | -- |
| APGAR 5 minute (IQR) | 8(8-9) | -- |
| Neonatal pneumonia, n(%) | 5(2.3) | 1.0-5.2 |
| Neonatal COVID-19, n(%) | 1(0.5) | 0.1-2.5 |
| Perinatal deaths, n(%) | 12(5.3) | 3.0-9.0 |
| Intrauterine fetal death, n(%) | 7(3.1) | 1.5-6.2 |
| Neonatal death, n(%) | 5(2.2) | 1.0-5.1 |

\* Neonates N = 227 (6 Twin, 7 fetal deaths)

In relation to births, a prematurity of 26.9% (59/221) was found, 12.2% (27/221) presented growth restriction or small for gestational age and perinatal mortality of 5.3% (12/227, includes twins), predominantly intra-uterine death.

Antiviral treatment was only offered to 4% of the cohort and of them 75% was post-birth, see Table 2. There was a need for mechanical ventilation in 5.9% (95% CI 3.6-9.6%) of the group and four of them died (1.6%), table 2.

## DISCUSSION

This is the first study from Panama and Central America and one of the few in Latin America on SARS-CoV-2 infection. The diagnosis of the infection is made predominantly in the third trimester, one in every 4 pregnant women has obesity as a risk factor, one third are asymptomatic, pre-eclampsia is the most frequent obstetric complication, one in ten presented the severe form of the disease and one in every 64 infected dies. There is a high frequency of prematurity, perinatal mortality is also high, and neonatal infection is rare.

Obesity is a known risk factor ^10^ for becoming infected with SARS-CoV-2 and this finding has also been reported in other studies of pregnant women ^11-12^; we found that obesity is present as a risk factor in 24.9% of pregnant women and the average body mass index in the entire population is in overweight values (28.4 kg / m2).

Another very interesting finding in our cohort is the gestational age at the time of making the diagnosis, 89% were more than 27 weeks of gestation; This finding is also reported in previous studies ^11-14^; a greater possibility of maternal infection in the third trimester is evident. This is a finding that deserves a follow-up study to assess prognosis and complications according to the trimester of infection. So far it has not been analyzed in large cohorts, despite the fact that there appears to be a greater susceptibility to infection in the third trimester.

Surprisingly, we found that pre-eclampsia with or without severe form and gestational hypertension were diagnosed in one of every 5 patients with COVID-19. This finding represents twice that diagnosed in a population before COVID-19 ^15,16^, in addition, this finding has not been reported in previous studies with COVID-19 ^11-14^. Definitely, more research is needed on the possible relationship, association or imitation between pre-eclampsia and maternal infection by SARS-CoV-2 ^17^, theoretically, it is possible to explain the greater presence of pre-eclampsia and gestational hypertension in pregnant women with COVID-19 ^18^.

The severity, need to admit to the intensive care unit, need for mechanical ventilation and lethality is definitely higher in pregnant women with COVID-19 ^5,19^. We found that the need to admit to intensive care units was 6.7%, being lower than that reported in the USA ^5^, but with a 5.9% possibility of endotracheal intubation which is double that reported in the USA, the lethality is similar to that reported in other latitudes ^5,12,14^, and possibly lower than in brazil ^19^.

Births before 37 weeks (prematurity) was found in 26.9% of the studied cohort, largely induced by the severity of the infection, and complications such as hypertension and premature rupture of membranes. Similar findings have been previously reported ^12,13,20^, however, it is more than twice the risk of that reported in the UK ^11^.

The perinatal mortality found in this study (5.3%) is very high compared to the UK, Wuhan and Sweden ^11,21^, and similar to that reported in a multinational study ^14^. The factor that contributes the most to mortality in our cohort is prematurity, since 67% (8/12) were less than 28 weeks old at the time of birth.

The limitations of this survey is the type of study (simple survey); the study does not involve the entire population; short duration, it was only for a period of 6 months, newborns are not followed-up after birth and do not know the risk of neonatal infections.

The main strengths are the contribution for the country and obvious for Latin America regarding the experience and results of mothers with COVID-19 and their perinatal outcomes, all patients have RT-PCR confirming the diagnosis of SARS-CoV-2.

In summary, this observational cohort study conducted in Panama with COVID-19 patients diagnosed with RT-PCR shows a high percentage of maternal complications, high admission to the ICU and mechanical ventilation, with about 2% maternal death; in addition to obstetric complications such as hypertension, premature rupture of membranes, high rate of prematurity and perinatal lethality. At the time of birth (using for PCR diagnosis) neonatal infection with SARS-CoV-2 is very low.

## Data Availability

all data is available

## AUTHORS CONTRIBUTIONS

All authors contributed equally for study conception and study design. JS, JE, LCC, AQ and PVD were responsible for data collection. JS, JE, CL, JNC, RdG, PVD and SCB were responsible for data analysis and interpretation. JS, JE, and PVD wrote the first draft of the paper. All authors reviewed and provided comments on the first draft and approved the final manuscript. The corresponding author attests that all listed authors meet the authorship criteria and that no others meeting the criteria have been omitted.

